# A Systematic Review and Dose-Response Meta-Analysis of the Association Between Nitrate & Nitrite intake and Gastroesophageal Cancer Risk: A Study Protocol

**DOI:** 10.1101/2023.12.01.23299297

**Authors:** Mohammadreza Ghasemi, Mohammad Bahrami koutenaei, Alireza Ghasemi, Reza Alizadeh-navaei, Mahmood Moosazadeh

## Abstract

**Objectives:** The global burden of cancer underscores the necessity for evidence-based strategies to identify and manage risk factors. Nitrate and nitrite are inherent byproducts resulting from microbial-mediated nitrogen oxidation in plants, soil, or water. Fruits, vegetables, and combinations thereof are integral constituents of a nutritious diet. Despite the potential carcinogenicity of N-nitroso compounds (NOCs), certain epidemiological investigations have reported a lack of correlation between the dietary intake of nitrate, nitrite, and NOCs and the occurrence of cancer in human subjects.

**Methods:** PubMed/MEDLINE, Embase, WoS (Clarivate Analytics), Proquest, Scopus, and Google Scholar as electronic databases will be precisely searched for observational studies that assessed the relationship between nitrate/nitrite levels and gastric/esophageal cancer. In this study, an assessment will be conducted on studies spanning January 1, 1980, to October 30, 2023. The inclusion criteria will not restrict language. Discrepancies among the reviewers at various stages of the study, including screening, selection, quality assessment, and data extraction, will be resolved through consensus. In instances where disagreements persist unresolved, a third reviewer will decide. The combination method will be employed based on methodological similarities in the chosen articles, utilizing either the Random Effect Model or the Fixed Effect Model. Additionally, forest plots will be generated for the included articles. Statistical heterogeneity will be evaluated using the I2 statistic and the Q-statistic test. Furthermore, funnel plots will be employed to assess non-significant study effects and potential reporting bias. Eggers and Beggs tests will be executed, and the identification of publication bias will rely on significant findings (P < 0.05).

**Conclusion:** The findings of this study should be of benefit to governments and researchers in maintaining safe levels of nitrate and nitrite in drinking water as well as preventing Gastroesophageal cancers.

## 1. Background

Gastric cancer (GC) manifests as a markedly aggressive and rapidly proliferating cancers. A considerable number of patients with gastric cancer exhibit partial metastasis at the time of diagnosis. Despite a decrease in the incidence of gastric cancer (GC) in recent decades, it maintains its position as the fifth most prevalent cancer, following lung, breast, colorectal, and prostate cancers.

Additionally, it stands as the third leading cause of cancer-related mortality globally, surpassed only by lung and colorectal cancers.(1)

Esophageal cancer poses a significant global health challenge. As per projections derived from the GLOBOCAN project, there were 572,034 newly diagnosed cases of esophageal cancer in 2018, accounting for 3.2% of all newly reported cancer cases worldwide. Additionally, 508,585 deaths were attributed to esophageal cancer in the same year.(2,3)

Given the escalating global burden of cancer, there is an escalating need for evidence-based approaches to identifying and managing risk factors associated with cancer development. While the precise causes of cancer remain incompletely understood, a multitude of factors, both non-modifiable (such as gender, age, and genetic factors) and modifiable (including diet and lifestyle factors), have been identified as contributors to increased risk. Notably, one-third of cancer-related deaths can be attributed to behavioral and dietary risks. In recent years, emerging evidence has instigated a paradigm shift in our comprehension of the impact of dietary nitrate and nitrite on human health, particularly concerning cancer risk.

Nitrate and nitrite are inherent byproducts resulting from the microbial-mediated oxidation of nitrogen in plants, soil, or water. Nitrate, being the predominant nitrogen source in soils, plays a crucial role in the production of amino acids in plants. It serves as a pivotal nutrient orchestrating optimal plant growth and developmental processes.(4)

Traditionally, elevated consumption of nitrate and nitrite was perceived as potentially harmful food additives and classified as likely human carcinogens, particularly in situations facilitating endogenous nitrosation and subsequent N-nitroso compound (NOC) formation. Presently, it is acknowledged that nitrate, nitrite, and nitrosamine naturally exist in fruits and vegetables, integral components of a healthful diet. This acknowledgment stems from compelling evidence supporting their substantial health benefits, particularly in the context of cancer prevention.(5–7)

The conversion of nitrogen compounds to nitrosamine has been linked to the increased risk of gastrointestinal cancer. Conversely, endogenously produced nitric oxide (NO) from naturally occurring nitrate in plants is implicated in blood pressure regulation, thereby contributing to enhanced cardiovascular health. The intake of inorganic nitrate positively influences endothelial function, leading to a significant reduction in cardiovascular disease (CVD) risk factors, including decreased platelet aggregation and arterial rigidity.(8–10)

Fruits, vegetables, and combinations thereof are recognized as integral constituents of a nutritious diet. The naturally occurring nitrate and nitrite found in these food sources are believed to confer advantageous effects in terms of cancer prevention and contribute to vascular and metabolic well-being. The nitrite–NO pathway has been demonstrated to exhibit antihypertensive properties.

Consequently, despite the substantial consumption of nitrate from vegetable sources, its intake is unlikely to be deemed problematic.(11)

Despite the potential carcinogenicity of N-nitroso compounds (NOCs), certain epidemiological investigations have reported a lack of correlation between the dietary intake of nitrate, nitrite, and NOCs and the occurrence of cancer in human subjects. Overall, epidemiologic studies that have examined associations between nitrate, nitrite, and NOC compounds and various types of cancer in humans have returned mixed and, in some cases, complex results.(12–14)

In this study, this systematic review aims to evaluate the association and clarify the relationship between dietary consumption of nitrate and nitrite and risk of gastric or esophageal cancer in humans. Furthermore, our objective is to elucidate the dose-response pattern of the relationship between nitrate/nitrite and gastric/esophageal cancer.

## 2. Objectives

### 2.1 Primary Objective

The main objective of this study is to examine the association between dietary nitrate intake levels and the gastric cancer.

### 2.2 Secondary Objectives

1. Assessing the relationship between levels of dietary nitrite intake and gastric cancer.
2. Assessing the relationship between levels of dietary nitrate intake and esophageal cancer.
3. Assessing the relationship between levels of dietary nitrite intake and esophageal cancer.
4. Assessing potential heterogeneity and finding its sources.

## 3. Methods

This systematic review and meta-analysis’s research protocol adhere to the guidelines outlined in the Cochrane Handbook, and it has been duly registered under the PROSPERO registration number CRD42023398071. The methodology for study selection will be articulated in accordance with the Preferred Reporting Items for Systematic Review and Meta-Analysis Protocols (PRISMA-P) 2015 guidelines.(15,16)

### 3.1 Patient and Public Involvement

No patients will be involved.

### 3.2 Inclusion and Exclusion Criteria

#### 3.2.1 Study Types

This study will include observational designs, including cross-sectional, case-control, and cohort studies, which investigate the association between nitrate and nitrite exposure and gastroesophageal cancer. Notably, case reports, case series, clinical trial studies, review articles, and protocol papers will be excluded.

#### 3.2.2 Types of Participants

People of all ages, with diagnosis of gastric or esophageal cancer wil be the Participants of this study.There will be no specific restrictions on age or sex. In vitro and in vivo studies will be excluded

### 3.3 Exposure

Included studies will have at least two levels of nitrate or nitrite exposure in diet. There is no limit to the units of reported nitrate or nitrite.

### 3.4 Control

Controls will refer to participants with gastric or esophageal cancer and normal levels of nitrate or nitrite in their diet.

### 3.5 Outcomes

The occurrence of gastric or esophageal cancer is the outcome of this study, which is considered a categorical variable. The effect size is the odds ratio (OR), which is defined as the odds of the occurance of gastric or esophageal cancer in people with high levels of nitrate or nitrite in diet compared to people who received normal levels of nitrate or nitrite.

### 3.6 Search Strategy Components

To attain a comprehensive search, the study will employ the PECO (Patient, Exposure, Comparison/Control, and Outcome) framework to formulate search terms pertaining to the exposure (nitrate or nitrite) and outcome (gastric or esophageal cancer) elements. The identified key search terms are delineated in Table 1, with appropriate formatting tailored to each database. Thesaurus systems incorporating Medical Subject Headings (MeSH) and Emtree, along with insights from experts, will be utilized to acquire equivalent terms for the components.

**Table 1.**
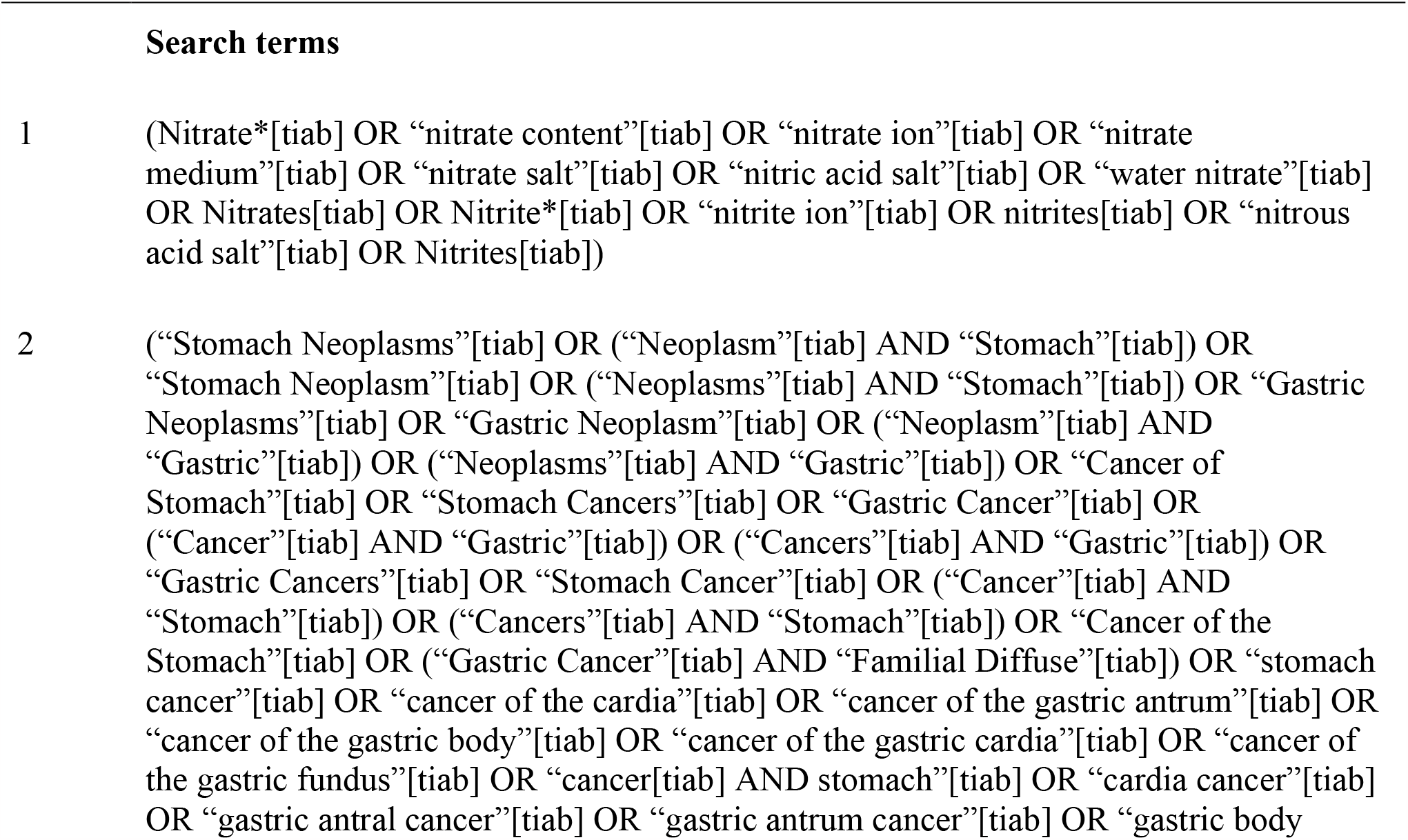
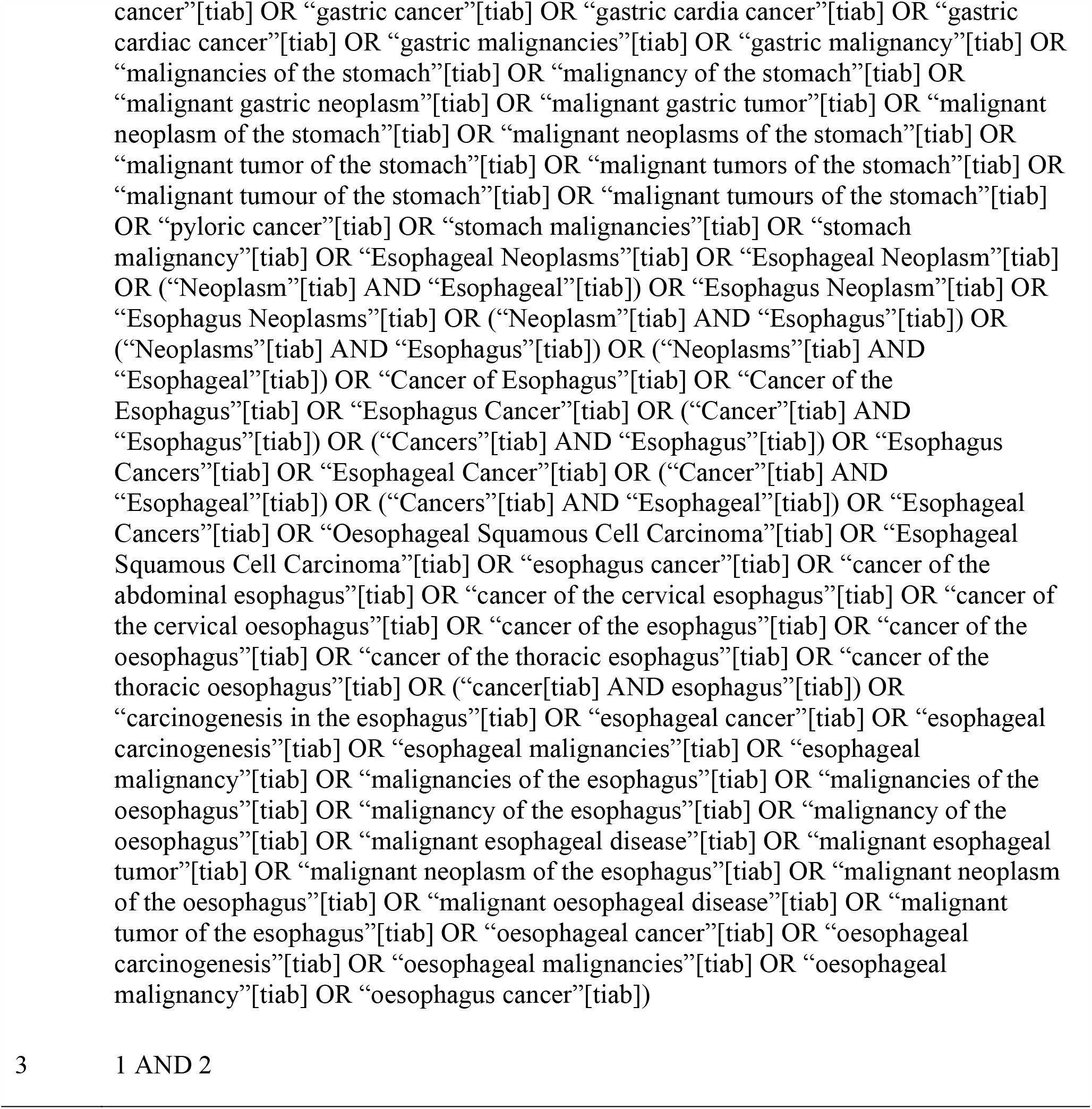
Search Strategies Used in PubMed/MEDLINE.

### 3.7 Electronic Database Search

Systematic searches will be executed using electronic databases, including PubMed/MEDLINE, Embase (Embase.com), Scopus, WoS (Clarivate Analytics), Google Scholar, and ProQuest.

### 3.8 Grey literature

Theses related to the subject will be identified through the electronic databases of Scopus and ProQuest. Additionally, electronic databases will be employed to locate conference papers and proceedings.

### 3.9 Publication date

This study will encompass all relevant research studies published within the timeframe from January 1, 1980, to October 30, 2023.

### 3.10 Publication language

In the present study, there would not be any language limitations. Any studies written in a language other than English, which are ultimately chosen for inclusion, must undergo an initial translation using Google Translate. Subsequently, they will be recheckd by an official translator.

### 3.11 Constructing the Search Syntax

As delineated in Table 1, the elements designated as ’Exposure (nitrate/nitrite)’ and ’Outcome (Gastric/Esophageal cancer)’ will be targeted for the identification of pertinent studies. The execution of a comprehensive search is anticipated to yield the maximal number of observational studies within electronic databases. The formulated search syntax is designed to be adaptable for application to various electronic databases. The detailed search syntax tailored for PubMed is outlined in Table 1. Subsequently, all searches conducted across diverse databases will be consolidated within Endnote software.

### 3.12 Study Screening and Selection

The search protocol will be implemented, adhering to the specified syntax tailored to each electronic database. One of the authors (MG) will undertake the evaluation of titles and abstracts in accordance with a predetermined checklist, according to the established inclusion criteria during the screening stage. Articles related to the subject of the study will be identified and extracted. studies failing to meet any of the inclusion criteria will be excluded at this stage. Papers lacking sufficient information on one or more inclusion criteria will be temporarily considered, with the final decision based on a comprehensive examination of the complete texts.

In the subsequent selection phase, two contributors (AG and MB) will meticulously assess the full texts of the studies identified during the screening stage, independently determining the conclusive selection of studies. Any discord arising during these phases will be resolved through consensus. In instances where consensus cannot be reached, an expert’s opinion (MM) will be sought for resolution.

### 3.13 Risk of bias Assessment

The evaluation of the methodological quality of studies will be conducted utilizing the Joanna Briggs Institute critical appraisal for cohort and case-controls (JBI checklist) (17). The comprehensive risk of bias in each included study will be categorized as “high,” “low,” or “unclear.”

This evaluation process will be independently performed by two authors (MG and AG). In the event of disagreements, resolution will be sought through consensus, and should consensus prove elusive, the perspective of a third expert (MM) will be solicited to arbitrate the matter.

### 3.14 Data extraction

Two authors, MG and MB, will complete the data extraction form for all the eligible papers, independently. Any disagreement in their assessments will be discussed to achieve a consensus, or alternatively, the viewpoint of a third expert (MM) will be sought for resolution.

The data extraction form will encompass the following information: the first author’s name, publication year, study country, study design, study duration, sample size, number of cases, quality scores of papers, cancer type, outcome type (occurrence/mortality), nitrate dose, nitrite, point estimate of odds ratio (OR) for each level of exposure, upper bound of the confidence interval of OR for each level of exposure, and lower bound of the confidence interval of OR for each level of exposure.

In instances where the included studies have incomplete data, efforts will be made to contact the authors for data retrieval. A paper will be excluded after three unsuccessful attempts to elicit a response from the study authors.

### 3.15 Data Synthesis and Analysis

Brief details pertaining to each study included in the analysis will be presented in a table. This table will encompass essential information such as the first author’s name, study country, study design, number of cases, nitrate/nitrite categories, reported odds ratio (OR) with 95% confidence interval (CI), and the specific cancer sites (stomach or esophageal).

### 3.16 Statistical Analysis

Stata V.14 software (StataCorp, USA) will be applied for the statistical analysis in this study.

### 3.17 Assessment of heterogeneity

To assess the statistical heterogeneity within the included studies, the I2 statistic and Q-statistic tests, along with their corresponding 95% Confidence Intervals (CIs), will be employed. Heterogeneity values categorized as 0%-40%, 30%-60%, 50%-90%, and 75%-100% will be interpreted as ’perhaps not important,’ ’moderate heterogeneity,’ ’substantial heterogeneity,’ and ’considerable heterogeneity,’ respectively. Regarding the Q-test, a significance level of P < 0.05 will be deemed statistically significant.

### 3.18 Assessment of Publication Bias

Conducting a comprehensive search at the initiation of the study constitutes the primary strategy to avoid publication bias. Additionally, funnel plots will be constructed to assess non-significant study effects and potential reporting bias. Egger’s test and Begg’s test will be conducted, and the identification of significant results (P < 0.05) may indicate the presence of publication bias.

Subsequently, the non-parametric ’trim and fill’ method will be implemented to rectify this form of bias.

### 3.19 Missing Data

Regarding probable missing data of the final papers, efforts will be made to obtain contact information for the corresponding authors in order to establish communication and acquire the missing data. Failure to establish contact with the authors will result in the exclusion of their studies.

## 4. Conclusions

The present systematic review and meta-analysis will assess the dose-response relationship between nitrate/nitrite and gastrointestinal cancer. Researchers and governments are expected to benefit from the results of this study since they will be able to use the findings of the study to control levels of nitrate/nitrite in dietary products and prevent gastric or esophageal cancer. The results of this study will be reported in publications and presentations at conferences.

## 5. Strengths and Limitations of this study

- This study will examine the association between nitrate/nitrite and gastric/esophageal cancer in a dose-response manner.
- The reporting of data will adhere to the Preferred Reporting Items for Systematic Reviews and Meta-Analysing Protocols.
- The study employs a comprehensive search strategy based on thesaurus systems, including Emtree and MeSH. It conducts searches across prominent databases such as Scopus, MEDLINE/PubMed, WOS, Embase, Google Scholar, and ProQuest, spanning an extensive time frame.
- Potential methodological biases in the primary studies included may introduce uncertainty into the findings of the present study.

## Data Availability

All data produced in the present study are available upon reasonable request to the authors

## 6. Acknowledgments

The authors wishes to acknowledge the Gastrointestitional Cancer Research Center, Mazandaran University of Medical Sciences for their support, which have been invalubale in the completion of this research.

## Conflict of Interest

The authors declare that they have no conflict of interest.

### Ethical Approval

Since patient information will be sourced from previously published papers, there is no requirement for seeking ethical approval. Any deviations from the established protocol will be thoroughly documented in the final report.

## References

1. Bray F, Ferlay J, Soerjomataram I, Siegel RL, Torre LA, Jemal A. Global cancer statistics 2018: GLOBOCAN estimates of incidence and mortality worldwide for 36 cancers in 185 countries. CA Cancer J Clin [Internet]. 2018 Nov;68(6):394–424. Available from: http://www.ncbi.nlm.nih.gov/pubmed/30207593

2. Thrift AP. Global burden and epidemiology of Barrett oesophagus and oesophageal cancer. Nat Rev Gastroenterol Hepatol. 2021;18(6):432–43.

3. Organization WH. Cancer Today: Data visualization tools for exploring the global cancer burden in 2020. 2020; Available from: https://gco.iarc.fr/today/home

4. Raddatz N, Morales de los Ríos L, Lindahl M, Quintero FJ, Pardo JM. Coordinated Transport of Nitrate, Potassium, and Sodium. Front Plant Sci [Internet]. 2020 Mar 6;11. Available from: https://www.frontiersin.org/article/10.3389/fpls.2020.00247/full

5. Jones RR, Weyer PJ, DellaValle CT, Inoue-Choi M, Anderson KE, Cantor KP, et al. Nitrate from Drinking Water and Diet and Bladder Cancer Among Postmenopausal Women in Iowa. Environ Health Perspect [Internet]. 2016 Nov;124(11):1751–8. Available from: http://www.ncbi.nlm.nih.gov/pubmed/27258851

6. Bradbury KE, Appleby PN, Key TJ. Fruit, vegetable, and fiber intake in relation to cancer risk: findings from the European Prospective Investigation into Cancer and Nutrition (EPIC). Am J Clin Nutr [Internet]. 2014 Jul;100 Suppl:394S–8S. Available from: http://www.ncbi.nlm.nih.gov/pubmed/24920034

7. Gonzalez CA, Lujan-Barroso L, Bueno-de-Mesquita HB, Jenab M, Duell EJ, Agudo A, et al. Fruit and vegetable intake and the risk of gastric adenocarcinoma: A reanalysis of the european prospective investigation into cancer and nutrition (EPIC-EURGAST) study after a longer follow-up. Int J Cancer [Internet]. 2012 Dec 15;131(12):2910–9. Available from: https://onlinelibrary.wiley.com/doi/10.1002/ijc.27565

8. van den Brand AD, Beukers M, Niekerk M, van Donkersgoed G, van der Aa M, van de Ven B, et al. Assessment of the combined nitrate and nitrite exposure from food and drinking water: application of uncertainty around the nitrate to nitrite conversion factor. Food Addit Contam Part A Chem Anal Control Expo Risk Assess [Internet]. 2020 Apr;37(4):568–82. Available from: http://www.ncbi.nlm.nih.gov/pubmed/31944907

9. Keller RM, Beaver L, Prater MC, Hord NG. Dietary Nitrate and Nitrite Concentrations in Food Patterns and Dietary Supplements. Nutr Today [Internet]. 2020 Sep;55(5):218–26. Available from: 10.1097/NT.0000000000000253

10. Karwowska M, Kononiuk A. Nitrates/Nitrites in Food—Risk for Nitrosative Stress and Benefits. Antioxidants [Internet]. 2020 Mar 16;9(3):241. Available from: https://www.mdpi.com/2076-3921/9/3/241

11. Salehzadeh H, Maleki A, Rezaee R, Shahmoradi B, Ponnet K. The nitrate content of fresh and cooked vegetables and their health-related risks. Sharma RR, editor. PLoS One [Internet]. 2020 Jan 9;15(1):e0227551. Available from: https://dx.plos.org/10.1371/journal.pone.0227551

12. Bryan NS, Alexander DD, Coughlin JR, Milkowski AL, Boffetta P. Ingested nitrate and nitrite and stomach cancer risk: An updated review. Food Chem Toxicol [Internet]. 2012 Oct;50(10):3646–65. Available from: https://linkinghub.elsevier.com/retrieve/pii/S0278691512005406

13. Espejo-Herrera N, Cantor KP, Malats N, Silverman DT, Tardón A, García-Closas R, et al. Nitrate in drinking water and bladder cancer risk in Spain. Environ Res [Internet]. 2015 Feb;137:299–307. Available from: https://linkinghub.elsevier.com/retrieve/pii/S0013935114003983

14. Lundberg JO, Weitzberg E, Gladwin MT. The nitrate–nitrite–nitric oxide pathway in physiology and therapeutics. Nat Rev Drug Discov [Internet]. 2008 Feb;7(2):156–67. Available from: https://www.nature.com/articles/nrd2466

15. Moher D, Liberati A, Tetzlaff J, Altman DG. Preferred Reporting Items for Systematic Reviews and Meta-Analyses: The PRISMA Statement. PLoS Med [Internet]. 2009 Jul 21;6(7):e1000097. Available from: https://dx.plos.org/10.1371/journal.pmed.1000097

16. Stroup DF. Meta-analysis of Observational Studies in Epidemiology<SUBTITLE>A Proposal for Reporting</SUBTITLE>. JAMA [Internet]. 2000 Apr 19;283(15):2008. Available from: http://jama.jamanetwork.com/article.aspx?doi=10.1001/jama.283.15.2008

17. Moola S, Munn Z, Tufanaru C, Aromataris E, Sears K, Sfetcu R, Currie M, Qureshi R, Mattis P, LisyK MP-F. Chapter 7: Systematic reviews of etiology and risk. Joanna Briggs Inst Rev Manual [Internet]. 2017; Available from: https://reviewersmanual.joannabriggs.org/

